# Anti-SARS-CoV-2 Antibodies Persist for up to 13 Months and Reduce Risk of Reinfection

**DOI:** 10.1101/2021.05.07.21256823

**Authors:** Floriane Gallais, Pierre Gantner, Timothée Bruel, Aurelie Velay, Delphine Planas, Marie-Josée Wendling, Sophie Bayer, Morgane Solis, Elodie Laugel, Nathalie Reix, Anne Schneider, Ludovic Glady, Baptiste Panaget, Nicolas Collongues, Marialuisa Partisani, Jean-Marc Lessinger, Arnaud Fontanet, David Rey, Yves Hansmann, Laurence Kling-Pillitteri, Olivier Schwartz, Jérome De Sèze, Nicolas Meyer, Maria Gonzalez, Catherine Schmidt-Mutter, Samira Fafi-Kremer

## Abstract

Assessment of the kinetics of SARS-CoV-2 antibodies is essential in predicting protection against reinfection and durability of vaccine protection. Here, we longitudinally measured Spike (S) and Nucleocapsid (N)-specific antibodies in 1,309 healthcare workers (HCWs), including 916 COVID-19 negative HCWs and 393 convalescent COVID-19 for up to 422 days post-symptom. From month (M)1 to M7-9 post-infection, SARS-CoV-2 antibodies decreased moderately in convalescent HCWs in a biphasic model, with men showing a slower decay of anti-N (p=0.02), and a faster decay of anti-S (p=0.0008) than women. At M11-13, anti-N dramatically decreased (half-life: 283 days) while anti-S stabilized (half-life: 725 days) at a median of 2.39 log Arbitrary Units (AU)/mL (Interquartile Range (IQR): 2.10 -2.75). Overall, 69 SARS-CoV-2 infections developed in the COVID-19 negative group (incidence of 12.22 per 100 person-years) versus one in the COVID-19 positive group (incidence of 0.40 per 100 person-years), indicating a relative reduction in the incidence of SARS-CoV-2 reinfection of 96.7% (p<0.0001). Correlation with live-virus neutralization assay revealed that variants D614G and B.1.1.7, but not B.1.351, were sensitive to anti-S antibodies at 2.3 log AU/mL, while IgG ≥ 3 log AU/mL neutralized all three variants. After SARS-CoV-2 vaccination, anti-S levels reached at least 3 logs regardless of pre-vaccination IgG levels, type of vaccine, and number of doses. Our study demonstrates a long-term persistence of anti-S IgG antibodies that may protect against reinfection. By significantly increasing cross-neutralizing antibody titers, a single-dose vaccination strengthens protection against escape mutants.

## Introduction

Since the beginning of the pandemic, the hypothesis of waning humoral immunity in COVID-19 convalescent patients has raised many concerns about the reliability of population-based seroprevalence studies, and more critically about long-term antibody protection against reinfection and by extension the durability of vaccine protection. COVID-19 leads to the development of protective neutralizing antibodies in the vast majority of cases (1-4). Several reports suggested a rapid decline of SARS-CoV-2 antibodies as early as 3 months after infection(3, 5), while others reported persistence of antibody responses up to five months(4, 6). A recent rigorous study investigating T and B cell responses in convalescent COVID-19 patients reported that substantial immune memory is generated after COVID-19, and 95% of subjects retained immune memory at *≈* 6 months after infection(7). Furthermore, the presence of SARS-CoV-2 anti-spike (S) and anti-nucleocapsid (N) IgG antibodies were associated with a reduced risk of SARS-CoV-2 reinfection up to 7 months after initial infection(8-10). The recent emergence of SARS-CoV-2 variants with high transmissibility such as variant B.1.1.7, or decreased susceptibility to antibodies such as variant B.1.351, has raised the question of whether antibodies still protect against reinfection. Data on persistence and long-term efficacy of the immune response are therefore of vital importance in understanding the overall evolution of the pandemic and post-pandemic dynamics, especially in the era of emerging variants(11-14).

Here, using validated serological assays (15-17) on a large cohort of healthcare workers (HCWs) who have recovered from mild COVID-19, we described the dynamics of SARS-CoV-2 humoral response up to one year after COVID-19, and analyzed the incidence of reinfection within this period. Second, we used the S-Fuse live-virus neutralization assay(18), to assess the sensitivity of infectious SARS-CoV-2 variants to anti-S antibodies before and after vaccination, several months after primary infection.

## Results

### Cohort characteristics

This study involved 4290 samples from 1,309 HCWs, including 393 convalescent COVID-19 (here called COVID-19 positive) and 916 COVID-19 negative HCWs (Figure 1). The COVID-19 positive HCWs included 345 with a history of positive SARS-CoV-2 RT-PCR and 48 with positive serology only. Both COVID-19 positive and COVID-19 negative cohorts included various professional groups (nurses, doctors, caregivers and administrative staff), both with a median age of 39 (Interquartile Range (IQR) 30-51 and 30-50, respectively), and a predominance of females (76.8% and 78.5%). In COVID-19 positive HCWs, a history of contact with a COVID-19 case was reported in 66% of participants. COVID-19 consistent symptoms were reported by 383 participants (97.5%), including 367 (93.4%) and 16 (4.1%) with mild or moderate disease, respectively (Table 1). No severe cases were reported. All COVID-19 positive participants were sampled at month one (M1) (median: 31 days post symptom onset (DSO); IQR: 24-38), 383 at M3-6 (median: 107 DSO; IQR: 92-131), 346 at M7-9 (median: 215 DSO; IQR: 195-243) and lastly 233 at M11-13 (median: 374 DSO; IQR: 347-396), of which 93 were vaccinated against SARS-CoV-2 before M11-13 sampling. Only one asymptomatic reinfection was reported after nine months in this cohort. Conversely, among the 916 COVID-19 negative HCW, 69 (7.5%) reported a SARS-CoV-2 infection including 49 with symptoms (8 before M3-6, 29 before M7-9 and 32 before M11-13). This was confirmed by a positive RT-PCR test and by seroconversion in 62% and 100% of these cases, respectively.

**Table 1.** Characteristics of the 393 COVID-19 positive healthcare workers.

|  | Total | Females | Males | P value |
| --- | --- | --- | --- | --- |
| Number of COVID-19 positive HCWs | 393 | 302 | 91 | NA |
| Age (years), median (IQR) | 39.0<br>(29.6-50.5) | 40.0<br>(30.1-51.1) | 34.3<br>(28.6-44.6) | 0.0965 |
| BMI (kg/m²), median (IQR) | 23.6<br>(21.3-26.9) | 23.6<br>(21.0-27.8) | 24.0<br>(21.9-25.6) | 0.8157 |
| Blood group |  |  |  |  |
| A, n (%) | 162 (41.2) | 125 (41.4) | 37 (40.7) | 0.2112 |
| B, n (%) | 33 (8.4) | 23 (7.6) | 10 (11.0) |  |
| AB, n (%) | 16 (4.1) | 14 (4.6) | 2 (2.2) |  |
| O, n (%) | 129 (32.8) | 108 (35.8) | 21 (23.1) |  |
| Rhesus negative, n (%) | 60 (15.3) | 50 (16.6) | 10 (11.0) | 0.2448 |
| Unknown, n (%) | 53 (13.5) | 32 (10.6) | 21 (23.1) | 0.0045 |
| COVID-19 history |  |  |  |  |
| Contact with COVID-19 case, n (%) | 259 (65.9) | 200 (66.2) | 59 (64.8) | 0.8021 |
| Previous positive SARS-CoV-2 RT-PCR, n (%) | 345 (87.8) | 263 (87.1) | 82 (90.1) | 0.5837 |
| COVID-19 symptoms, n (%) | 383 (97.4) | 294 (97.4) | 89 (97.8) | 1.0 |
| Known date of symptoms onset, n (%) | 378 (96.2) | 289 (95.7) | 89 (97.8) | 0.5359 |
| Hospitalization, n (%) | 16 (4.1) | 10 (3.3) | 6 (6.6) | 0.2211 |
| Serum collection |  |  |  |  |
| HCW sampled at M1, n (%) | 393 (100) | 302 (100) | 91 (100) | 0.8042 |
| HCW sampled at M3-6, n (%) | 383 (97.5) | 294 (97.4) | 89 (97.8) |  |
| HCW sampled at M7-9, n (%) | 346 (88.0) | 275 (91.1) | 71 (78.0) |  |
| HCW sampled at M11-13, n (%) | 233 (59.3) | 181 (59.9) | 52 (57.1) |  |
| Time from symptom onset to serum collection at: |  |  |  |  |
| M1 (days), median (IQR; range) | 31 (24-38;<br>6-58) | 32 (24-38;<br>6-53) | 29 (24-37;<br>13-58) | 0.2087 |
| M3-6 (days), median (IQR; range) | 107 (92-131;<br>78-172) | 107.5 (92.3-131; 78-172) | 105 (90-130;<br>78-164) | 0.6583 |
| M7-9 (days), median (IQR; range) | 215 (195-243;<br>161-284) | 217 (196-246;<br>161-284) | 210 (194-237;<br>169-281) | 0.3388 |
| M11-13 (days), median (IQR; range) | 373 (347-396;<br>321-422) | 369 (346-396;<br>321-421) | 384 (348-396;<br>332-422) | 0.2053 |
| Vaccination |  |  |  |  |
| Single-dose vaccination between M7-9 and M11-13, n (% of M11-13) | 59 (25.3) | 43 (23.8) | 16 (30.8) | 0.3655 |
| Double dose vaccination between M7-9 and M11-13, n (% of M11-13) | 34 (14.6) | 27 (14.9) | 7 (13.5) | 1.0 |

BMI: body mass index; HCWs: Healthcare workers; IQR: Interquartile range; NA: not applicable; NS: not significant; RT-PCR: Real-time reverse transcriptase PCR. P values were calculated with Mann-Whitney, Chi-square and Fisher exact tests using the Graphpad Prism version 9.0.0 software.

**Figure 1:**
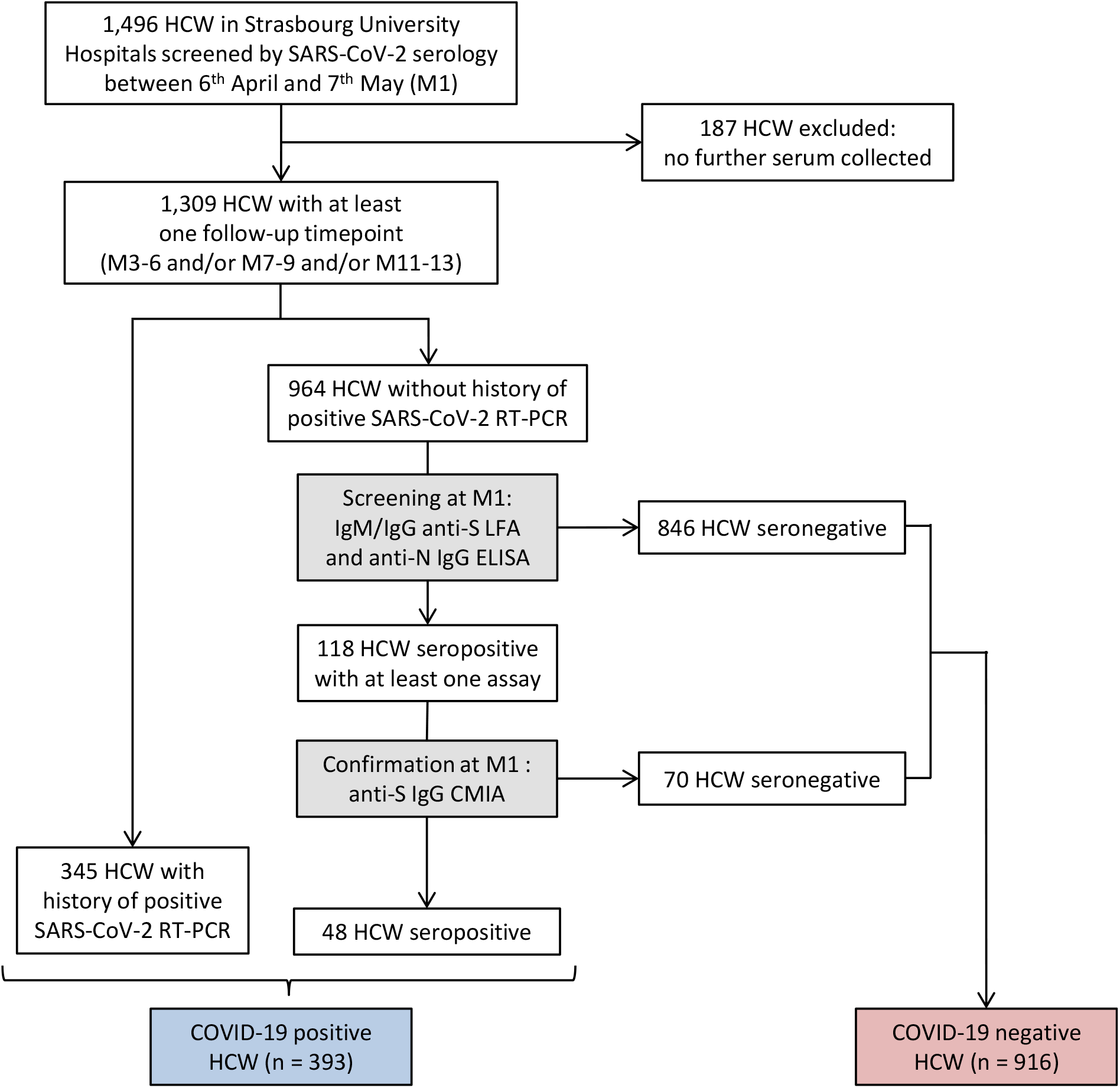
Flow chart of survey recruitment and serum sampling among the healthcare workers (HCWs) at Strasbourg University Hospital. First line serological screening was performed using two commercial assays: the Biosynex^®^ (COVID-19 BSS IgG/IgM) Lateral Flow Assay (LFA) detecting anti-spike (S) antibodies and the EDI™ Novel coronavirus COVID-19 IgG ELISA assay detecting the anti-nucleocapsid protein (N) IgG. A third assay, the Abbott SARS-CoV-2 IgG II Quant assay, measuring the anti-S IgG, was used to confirm seropositive samples. Serological testing on the first serum sample of each participant between 6 April and 7 May 2020 (M1) and on further sera collected at M3-6, M7-9, and M11-13 led to the establishment of two separate cohorts of COVID-19 positive or negative HCWs, both with serological follow-up. Anti-S: anti-spike protein; Anti-N: anti-nucleocapsid protein; CMIA: chemiluminescent microparticle immunoassay; ELISA: enzyme-linked immunosorbent assay; HCW: healthcare workers; LFA: lateral flow assay; RT-PCR: real-time reverse transcriptase PCR.

### Natural history of humoral response up to one year after COVID-19

We first sought to analyze the dynamics of SARS-CoV-2 humoral responses and its determinants in the aftermath of COVID-19 (e.g. natural history after primary infection, in the absence of vaccination). Seropositivity rates differ widely depending on: (1) isotypes (IgM or IgG), (2) antibody targets (N or S), (3) assays and (4) timepoints of serum collection (Figure 2A). At M1, the proportions of COVID-19 positive HCWs who tested positive for anti-S IgM and IgG using lateral flow assay (LFA) were 91.3% and 83.7%, respectively. Approximately half of individuals still had detectable antibodies at M11-13 (51.8% IgM and 56.8% IgG), showing a significant decrease in the rates of LFA-detected antibodies one-year after COVID-19 (both p<0.0001). Positivity rates of anti-N IgG response as measured by ELISA also significantly decreased from M1 (85.0%) to M11-13 (20.1%) (p<0.0001). Conversely, anti-S IgG response was maintained over time as shown by the persistence of seropositivity chemiluminescence microparticle immunoassay (CMIA)-based rates: 97.1% both at M1 and M11-13 (p=0.76).

**Figure 2:**
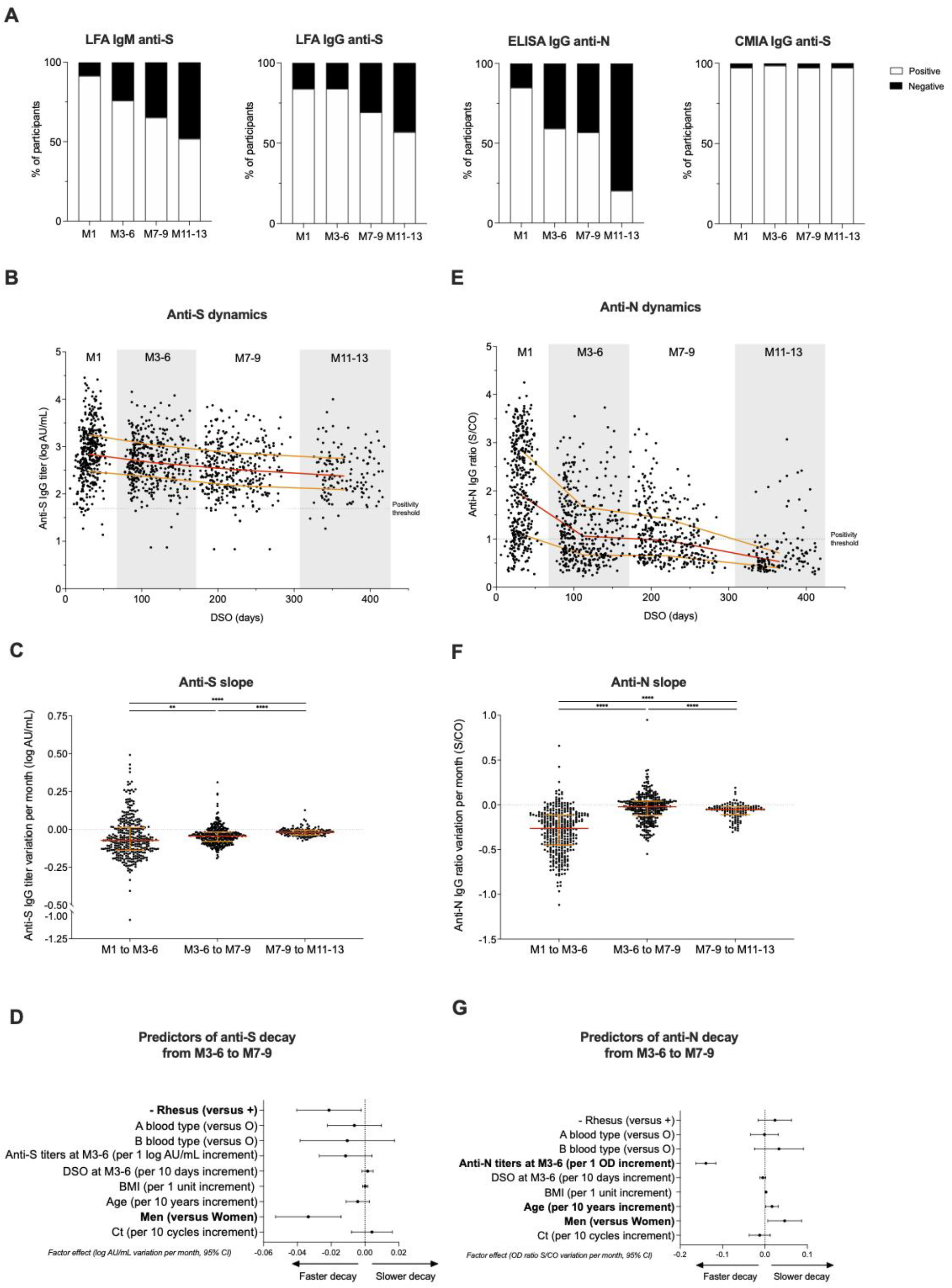
Dynamics and determinants of SARS-CoV-2 humoral responses after COVID-19. (**A**) SARS-CoV-2 seropositive rate over time among participants (M1 (n=393), M3-6 (n=383), M7-9 (n=346), M11-13 (n=139)) according to the different serological assays. (**B**) Dynamics of anti-S IgG titers expressed in log AU/mL (Abbott SARS-CoV-2 IgG II Quant assay) over time among HCWs with known first date of symptoms (M1 (n=369), M3-6 (n= 369), M7-9 (n= 332), M11-13 (n=137). (**C**) Variation of anti-S IgG titers per month expressed in log AU/mL between each time point (M1 to M3-6 (n=374), M3-6 to M7-9 (n=337), M7-9 to M11-13 (n=128)). (**D**) Associations between anti-S IgG titers decay between M3-6 to M7-9 and demographical, biological, and virological data and time of sampling at M3-6 expressed in days post symptoms onset (DSO) (n=337). (**E**) Dynamics of anti-N IgG expressed in ratio optical density (OD) signal/Cut Off (S/CO) (EDI™) over time among HCWs with known first date of symptoms (M1 (n=378), M3-6 (n= 369), M7-9 (n= 332), M11-13 (n=137)). (**F**) Anti-N IgG ratios variation per month expressed in ratio OD Sample/CO between each timepoints (M1 to M3-6 (n=383), M3-6 to M7-9 (n=337), M7-9 to M11-13 (n=128)). (**G**) Associations between anti-N IgG ratio decay between M3-6 to M7-9 and demographical, biological, and virological data and time of serum sampling at M3-6. For panel **B, C, E and F**, red lines represent median values and yellow lines the interquartile range for each population. \**p* value < 0.05; \*\**p* value < 0.01; \*\*\**p* value < 0.001, \*\*\*\**p* value <0.0001; calculated with non-parametric Wilcoxon paired tests using the Graphpad Prism version 9.0.0 software. For panel **D and G**, data are depicted as factor effects in multivariate linear regression, with a 95% confidence interval given (95%CI). Multivariate analyses were performed with R software version 4.0.3 (R Foundation for Statistical Computing, Vienna, Austria).

Next, we analyzed the dynamics of anti-S and anti-N IgG titers over time. Anti-S titers assessed by CMIA significantly decayed by 0.07, 0.04 and 0.02 log Arbitrary Units (AU/mL) per month from M1 to M3-6, M3-6 to M7-9 and M7-9 to M11-13, respectively (all p<0.01) (Figure 2B, C). The estimated half-life (t_1/2_) of each phase was of 202, 306 and 725 days, respectively. At M11-13, the median titer of anti-S IgG was 2.39 log AU/mL (IQR: 2.10-2.75), with 81.3% of participants showing IgG> 2.0 log AU/mL and 55.4% > 2.3 log AU/mL.

Next, we investigated the effect of age, sex, body mass index (BMI), blood group, rhesus status, DSO, and initial Ct values obtained by real-time reverse transcriptase PCR (RT-PCR) in nasal swabs, on CMIA anti-S IgG titers at M7-9 and on decay speed between M3-6 and M7-9 by univariate (not shown) and multivariate analyses. No significant difference in the absolute values of anti-S IgG titers was found in univariate analysis according to sex. However, antibody titers declined faster in men in univariate analysis between M3-6 and M7-9. By multivariate analysis (Figure 2D), anti-S IgG titers also decayed faster in men than in women with an acceleration in this decrease of - 0.033 log AU/mL per month (95% confidence interval 95%CI: - 0.053 to - 0.014; p=0.0008). Another factor significantly associated with faster decay was the rhesus-negative (Rh-) status, impacting decay by a factor of -0.021 log AU/mL per month (95%CI: - 0.002 to - 0.040; p=0.0008). Notably, no significant effect of age, BMI, blood group, DSO or initial Ct values on the anti-S titer slope was observed (Figure 2D).

Regarding ELISA anti-N IgG, a significant decay of ratios (optical density signal /Cut-Off: S/CO) per month was observed between the four study visits (Figure 2E). Interestingly, triphasic kinetic dynamics of anti-N IgG ratio over time was observed with an initial steep decay between M1 and M3-6 (median: -0.26 S/CO per month), followed by a slower decay up to M7-9 (−0.02) before a second drop up to M11- 13 (−0.05; all p<0.0001) (Figure 2F). The t_1/2_ of each phase was therefore 58, 682 and 283 days, respectively. This pattern differed from those of anti-S IgG titers. Univariate and multivariate analyses were conducted for anti-N ratio similarly to anti-S titers in order to identify potential predictor factors of anti-N IgG dynamics. Higher antibody ratios were found in men in univariate analysis at M7-9 compared to women. Moreover, a slower decay from M3-6 to M7-9 was revealed by multivariate analysis in men (0.046 S/CO per month; 95%CI 0.007-0.087; p=0.02) and in older participants (0.017 per 10-year age; 95%CI 0.002-0.032; p=0.03) (Figure 2G). Thus, male participants displayed a faster decay of anti-S antibodies and, conversely, a slower decay of anti-N antibodies.

We then assessed the relative incidence of SARS-CoV-2 infection in COVID-19 positive and COVID-19 negative HCWs during follow-up. Overall, 70 SARS-CoV-2 infections developed after enrollment: 1 in the COVID-19 positive group (incidence of 0.40 per 100 person-years) and 69 in the COVID-19 negative group (incidence of 12.22 per 100 person-years), indicating a relative reduction in the incidence of SARS-CoV-2 reinfection in the previously infected group of 96.7% (p<0.0001, Figure 3A,B). The only case of reinfection occurred in a 23 year old female medical student. She first developed a symptomatic, mild COVID-19 in March 2020 with a high viral load, identified by nasopharyngeal swab (Ct=17), leading to an anti-S and anti-N IgG seroconversion (2.6 log AU/mL and 1.0 OD S/CO after 96 DSO, respectively). The second episode in January 2021 was asymptomatic and revealed by a low viral load (Ct=34), detected six days after non-professional COVID-19 exposure. The reinfection was associated with positive anti-S IgM and a rebound of both anti-S IgG titer (3.6 log AU/mL) and anti-N IgG ratio (1.7 S/CO) without vaccination 22 days after a second positive RT-PCR. Altogether, our findings indicate that although anti-SARS-CoV-2 antibody titers do indeed decline, the risk of reinfection within a year post-infection remains low.

**Figure 3.**
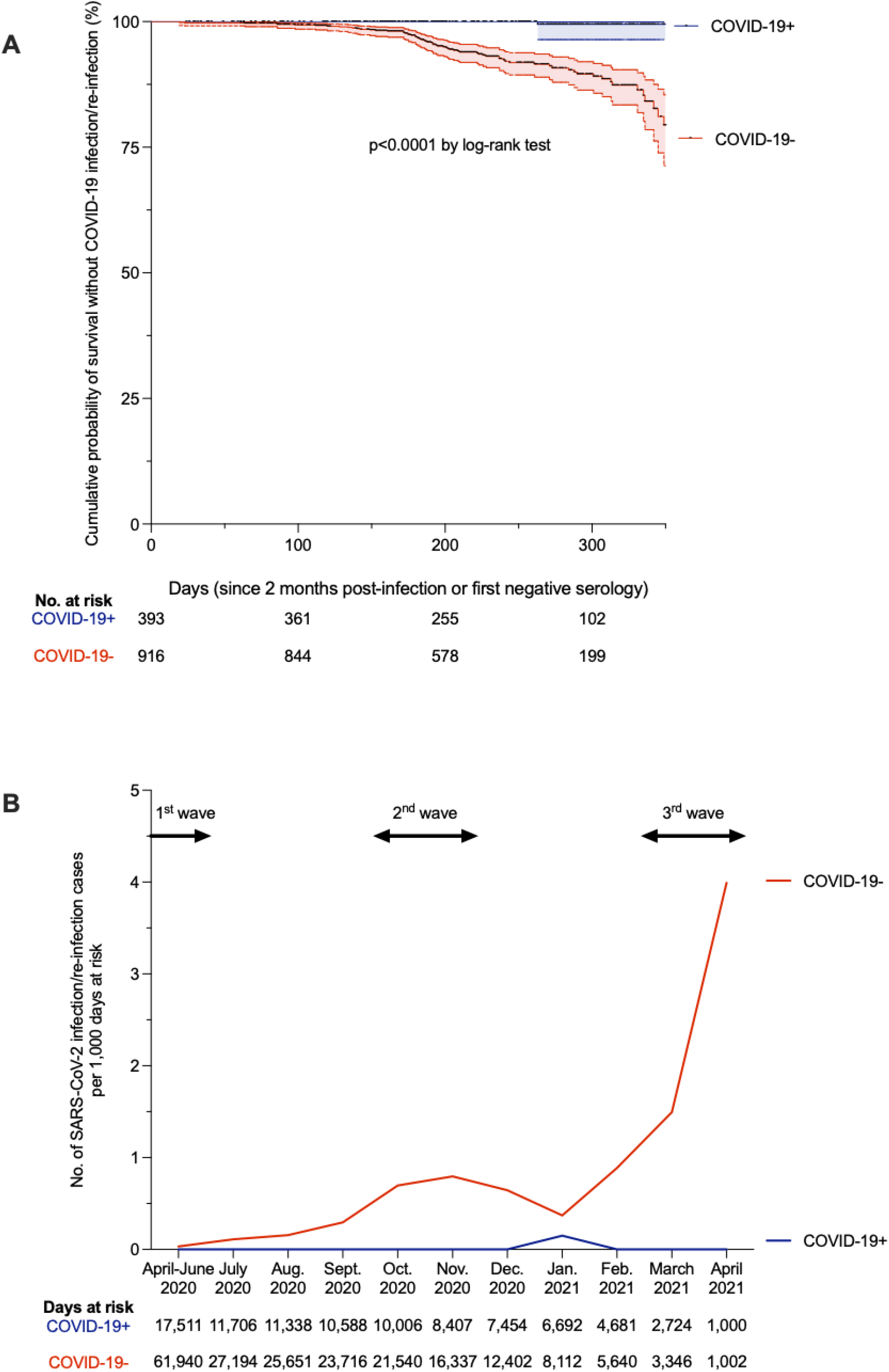
The risk of reinfection after a first COVID-19 episode. **(A)** Kaplan-Meier estimates of the probability of SRAS-CoV-2 infection protection. The cumulative probabilities of remaining free of SARS-CoV-2 infection among COVID-19 negative (COVID-19-) participants (red curve) and reinfection among former COVID-19 positive (COVID-19+) participants (blue curve) are shown on one year of follow-up (with 95% confidence interval, dotted lines). Exposition starts since the first negative serology for the COVID-19-group and since two months after initial SARS-CoV-2 infection for the COVID-19+ group, as described previously(10). SARS-CoV-2 infection/reinfection was assessed either by RT-PCR or/and serology. Vaccinated individuals were censored at the time of the first vaccine dose. The number of exposed participants is defined under the x axis. Comparison of survival curves was performed using log-rank test. Comparison and p value were computed using the Graphpad Prism version 9.0.0 software. **(B)** Calculated incidence of SARS-CoV-2 infection/reinfection per month, according to at-risk days during the follow-up of COVID-19+ (blue curve) and COVID-19-(red curve) individuals. Exposition starts since the first negative serology for the COVID-19-group and since two months after initial SARS-CoV-2 infection for the COVID-19+ group, as described previously(10). SARS-CoV-2 infection/reinfection was assessed either by RT-PCR or/and serology. Vaccinated individuals were censored at the time of the first vaccine dose. Data are represented according to calendar months to allow the reflection with national epidemic dynamics (epidemic waves depicted at the top of the graph with arrows). The number of at-risk days is shown under the x axis.

### Impact of SARS-CoV-2 vaccination on humoral response in COVID-19 positive HCWs

To investigate how SARS-CoV-2 antibodies evolve after COVID-19 vaccination in COVID-19 positive HCWs, serological results of the 93 COVID-19 positive HCWs who received at least one dose of vaccine between M7-9 and M11-13 visits were compared to those of the 139 unvaccinated participants with a M11-13 follow-up. Among vaccinated-participants, 59 received a single-dose from 1 to 99 days before M11-13 sampling, including 27 HCWs vaccinated with ChAdOx1 nCoV-19 vaccine (AstraZeneca), 4 with mRNA-1273 vaccine (Moderna) and 28 with BNT162b2 vaccine (Pfizer-BioNTech). The 34 other participants received two doses of BNT162b2 vaccine (n=33) or mRNA-1273 vaccine (n=1) and their M11-13 sera were collected from 3 to 94 days after the second dose of vaccine.

Six out of the seven participants sampled earlier than 6 days after a single dose vaccination still displayed anti-S antibody titers under 3 log AU/mL at M11-13 (Figure 4A). Conversely, a rebound of anti-S IgG titers was observed in all the 86 samples collected at least 6 days after vaccination with a median increase of 1.80 log AU/mL between M7-9 and M11-13 (IQR, 1.38 to 2.17; p<0.0001). Indeed, post-vaccination titers reached at least double the values measured at M1 post-infection. Antibody titers over 4 log AU/mL were found in 76 of these 86 vaccinated HCWs (88.4%) (Figure 4A, B). Altogether, our findings suggest that a strong humoral response is rapidly re-mobilized after a single-dose vaccination among COVID-19 positive individuals.

**Figure 4:**
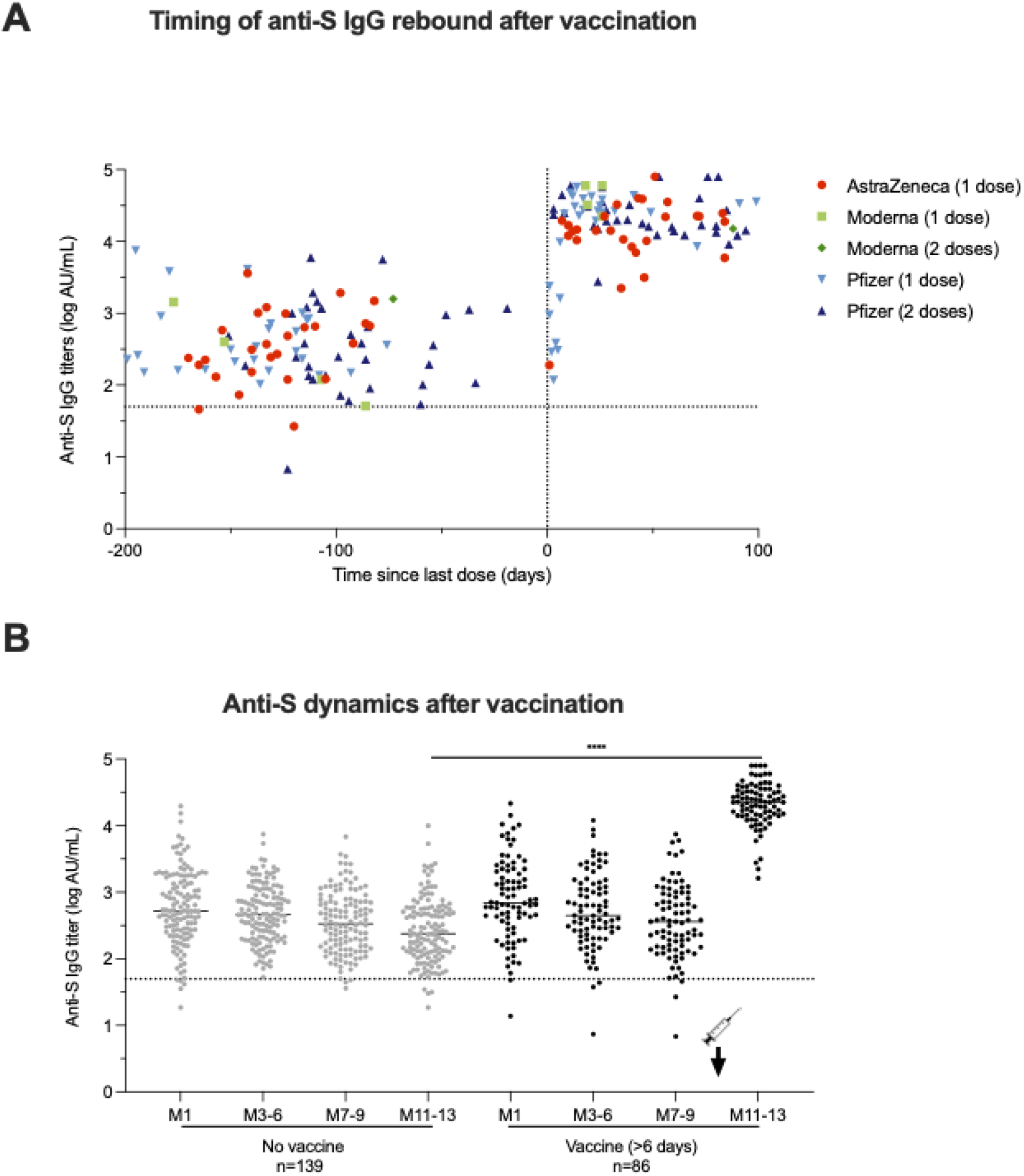
Impact of SARS-CoV-2 vaccination on humoral response in COVID-19+ HCWs. (**A**) Timing of the rebound in anti-S IgG titers following vaccination among the 93 COVID-19 positive HCWs who received at least one dose of vaccine against SARS-CoV-2 between M7-9 and M11-13 visits. The dotted vertical black line corresponds to the day of first injection (D0). Anti-S IgG titers among HCWs vaccinated with one dose of AstraZeneca vaccine are depicted in red circles, with one or two doses of Moderna vaccine in light and dark green squares, with one or two doses of Pfizer vaccine in light blue upwards and dark blue downwards triangles, respectively. (**B**) Comparison of anti-S IgG titer dynamics over time between 139 unvaccinated HCWs in grey dots and 86 HCWs vaccinated for more at least six days in black dots. *p value < 0.05; **p value < 0.01; ***p value < 0.001, ****p value <0.0001; calculated with non-parametric Wilcoxon paired tests and Mann-Whiney tests using the Graphpad Prism version 9.0.0 software. The dotted horizontal black line corresponds to the anti-S IgG titer positivity threshold.

### Sensitivity of infectious SARS-CoV-2 variants to anti-S antibodies at M11-13

To assess whether SARS-CoV-2 variants are sensitive to anti-S antibodies that persist at M11-13 with or without prior vaccination, sera collected at M11-13 from 28 COVID-19 positive HCWs (13 vaccinated and 15 unvaccinated) were analyzed with the S-Fuse live-virus neutralization assay(18) (Figure 5A). The 13 vaccinated HCWs had received a single dose, including 8 with ChAdOx1 nCoV-19 vaccine (AstraZeneca), 3 with BNT162b2 vaccine (Pfizer-BioNTech) and 2 with mRNA-1273 vaccine (Moderna). Sera collected from unvaccinated participants showed median neutralizing antibody titers of 2.31 log IC_50_ (IQR: 2.03-2.76), 2.10 log IC_50_ (IQR: 1.76-2.45) and 1.51 log IC_50_ (IQR: 1.48-1.87) against D614G, B.1.1.7 and B.1.351 live-strains, respectively. Sera from vaccinated participants showed a median neutralizing antibody titer of 4.01 log IC_50_ (IQR: 3.88-4.35), 4.03 log IC_50_ (IQR: 3.85-4.23) and 3.14 log IC_50_ (IQR: 2.99-3.58) against the same viral strains, respectively (Figure 5B). Strong correlation was observed at M11-13 between neutralizing antibody titers assessed by S-Fuse neutralization assay and anti-S IgG titers measured by CMIA with Spearman correlation coefficients of 0.934, 0.952 and 0.967 for variants D614G, B.1.1.7 and B.1.351, respectively (*p* values<0.0001) (Figure 5C). Anti-S titers around 2.3 log AU/mL neutralized D614G, B.1.1.7 but not B.1.351 variants at more than 2 log IC_50_. Anti-S IgG titers > 3 log AU/mL neutralized D614G, B.1.1.7 at > 2.5 log IC_50_ and B.1.351 at ≥ 2 log IC_50_. These anti-S IgG titers were reached by all vaccinated HCW regardless of pre-vaccination anti-S IgG titers, type of vaccine or number of vaccine doses. Based on the strong correlation between CMIA and neutralization assays, neutralizing titers were extrapolated to the remaining 124 unvaccinated HCWs and the 73 HCWs vaccinated (those with serum collected at least 6 days post-vaccination) to predict the strength of neutralization at M11-13 in all participants (Figure 5D). Altogether, our findings suggest that former COVID-19 positive individuals benefit from a single-dose vaccine and are able to efficiently neutralize current SARS-CoV-2 variants.

**Figure 5:**
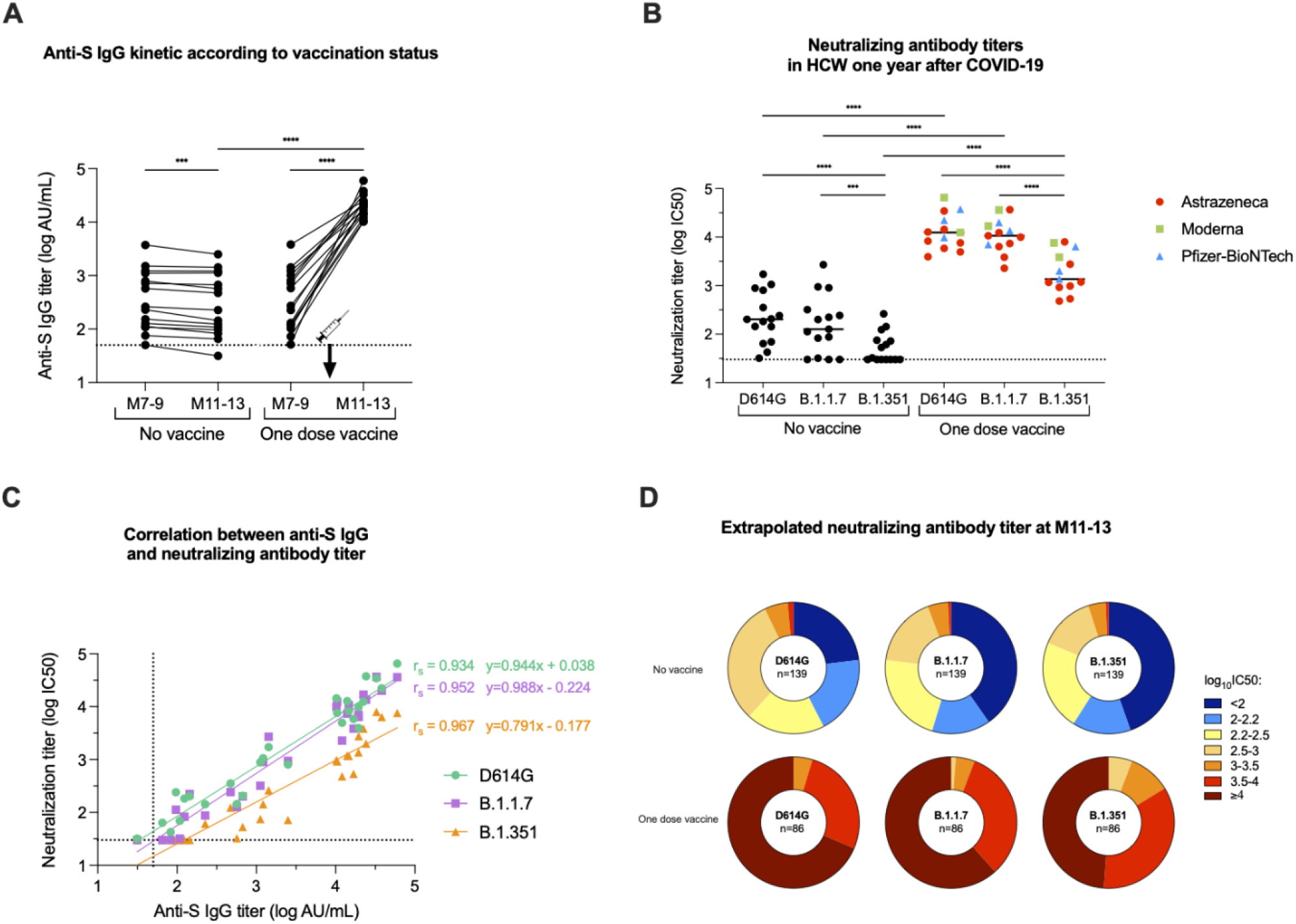
Neutralization efficiency after single-dose vaccination among the COVID-19+ HCWs. Neutralizing antibody titers against live-strains of D614G, B.1.1.7, and B.1.351 variants of SARS-CoV-2 were measured in sera collected at M11-13 for 13 single-dose vaccinated HCWs and 15 unvaccinated HCWs. (**A**) Anti-S IgG (log AU/mL) kinetics between M7-9 and M11-13 according to vaccination status. (**B**) Neutralizing antibody titers (log IC50) against the D614G, B.1.1.7, and B.1.351 variants measured at M11-13. The dotted black line corresponds to positivity threshold of neutralization assay. Neutralizing antibody titers measured in HCWs vaccinated with one dose of AstraZeneca vaccine (red circles), Moderna vaccine (green squares) or Pfizer-BioNtech vaccine(light blue triangles). (**C**) Spearman correlation between anti-S IgG titers (log AU/mL) and neutralizing antibody titer (log IC50) against D614G (green circles), B.1.1.7 (violet squares), and B.1.351 (orange triangles) variants measured at M11-13 in vaccinated (n=13) and unvaccinated (n=15) HCWs. The calculated correlation coefficients (r) and linear regression equations are depicted. (**D**) Pie charts depicting the frequency of log IC_50_ neutralization titer categories (extrapolated from CMIA anti-S titers) for all participants at M11-13 of according to the viral strain and the vaccination status. The number of participants is included at the center of the pie. *p value < 0.05; **p value < 0.01; ***p value < 0.001, ****p value <0.0001; calculated with non-parametric Wilcoxon paired tests or Spearman correlation. P values and correlation coefficients computed using the Graphpad Prism version 9.0.0 software.

## Discussion

The duration and effectiveness of adaptive immunity directed against SARS-CoV-2 after primary infection are key questions in understanding the coronavirus disease 2019 (COVID-19) pandemic. The present study involving a large cohort of HCWs followed prospectively over one year provides, for the first time, crucial information on persistence of circulating SARS-CoV-2 antibodies after mild COVID-19. We demonstrate that: i) anti-SARS-CoV-2 antibody titers evolve differently in men and women; ii) anti-S IgG stabilize at a median titer of 2.39 log AU/mL (IQR: 2.10 – 2.75) one year after symptom onset; iii) the risk of SARS-CoV-2 reinfection was reduced by 96.7% in the ensuing 12 months, iv)CMIA anti-S IgG titers strongly correlate with neutralization titers, v) Anti-S IgG titers around 2.3 log AU/mL efficiently neutralize D614G, B.1.1.7 but not B.1.351; vi) SARS-CoV-2 vaccination significantly increases anti-S antibodies to levels that neutralize all three variants regardless of pre-vaccination IgG levels, type of vaccine, or number of doses.

Our longitudinal study covered a serological monitoring of convalescent COVID-19 up to 422 days post-symptom, and showed that almost all COVID-19 positive (96%) still present detectable anti-S IgG one year after infection. A previous longitudinal study investigating anti-S IgG found relatively stable antibody titers over eight months after COVID-19(7). However, this study had data at only two time points and was not able to define a model for the kinetic of antibodies(7). In our study, follow-up at M1, M3-M6, M7-M9 and M11-13 showed a tri-phasic decay of anti-S antibodies. This segmented anti-S decay could reflect B cells turnover after infection(2). Although antibody titers were variable between unvaccinated convalescents, 81.3% retain anti-S IgG titers up to 2 log AU/mL and 55.4% up to 2.3 log AU/mL at M11-13. According to the correlation curve, titers over 2.3 log AU/mL are allowed to neutralize D614G and B.1.1.7 variants but less B.1.351, suggesting that most COVID-19 positive patients may be protected from reinfection by the former variants for at least one year after primary infection. It should be noted that our hospital faced three waves of COVID-19, from March – June 2020, September 2020 – January 2021 and from March 2021 to presently, with the current wave due to the B1.1.7 variant. During the period April 2020 – April 2021, 69 new infections were reported in COVID-19-negative participants while only one case of asymptomatic reinfection was reported in the COVID-19-positive participants. Although antibodies represent only a part of the immune response, this strongly suggests that COVID-19 positive patients develop a robust humoral immune response that reduces the risk of SARS-CoV-2 reinfection within at least one year.

Interestingly, all individuals who received a SARS-COV-2 vaccine displayed high antibody titers able to neutralize all three variants tested, regardless of pre-vaccine anti-S IgG levels, type of vaccine (mRNA or adenovirus-vector vaccines) or number of vaccine doses. The increase in antibody titer was observed as early as 6 days after vaccination. This suggests that a robust memory B cell response is established in COVID-19 convalescents, including those with low antibody titers. This is in line with the study of Dan *et al*. who performed an extensive characterization of memory B cells, revealing that the slight antibody decline occurring in convalescent individuals does not reflect a real waning of humoral immunity, but rather a contraction of the immune response, whilst antibody affinity maturation occurs, and anti-S memory B cells persist(7). Very recently, Wang et al reported that memory B cell clones expressing broad and potent anti-S antibodies are selectively retained in the repertoire at least one year after infection and expand after vaccination(19). These observations are very hopeful regarding the durability of humoral responses developed after COVID-19 and suggest that this protection against SARS-CoV-2 infection may last for years(19, 20).

Unlike anti-S antibody titers which stabilize over time, we observed a steep decay of anti-N IgG titers after seven to nine months post-infection, with only 20% of COVID-19 positive HCWs remaining seropositive after one year. Previous studies with a shorter monitoring period after infection found discrepant results regarding anti-N IgG persistence, depending on the commercial assay used. One study described a sustained humoral response up to ten months after infection(21), whereas another reported a significant decrease rapidly after infection, in line with our findings(22). These differences could be explained by increased avidity or affinity that compensates antibody loss, or by changes in recognized epitopes over time(22). Overall, our study show that serological assays targeting nucleocapsid should not be used preferentially over seroprevalence studies, even if they have the advantage of differentiating between natural infection and post-vaccinal immunity.

We evaluated several host factors as potential predictors of antibody titers, and of their kinetics up to 7 - 9 months after primary infection. While no differences in SARS-CoV-2 IgG titers were observed, their kinetics were influenced by sex and rhesus factor. Notably, men displayed a significantly faster decay of anti-S IgG and conversely, a significantly slower decrease of anti-N IgG titers between M3-6 and M7-9 after infection, independent of age and of titers measured at M3-6. Sex differences in the SARS-CoV-2 immune response were previously described early after infection. Takahashi and colleagues reported that female patients had more robust T cell activation than male patients in the early phase of SARS-CoV-2 infection(23). Other studies reported a higher peak of anti-S antibody titers in men early after infection followed by a steeper decay compared to females(7, 24-26). The initial, greater humoral response in convalescent men has been linked to the higher risk of severe disease in this population and to prolonged virus shedding(27, 28). However, this sex difference was also observed independently of age, severity of symptoms, or duration of symptoms(7, 24). We showed that differences in antibody kinetics depending on sex were still observed later than six months, independently of case severity since only mild cases, and a few moderate and asymptomatic cases, were monitored in our study. The sex differences in immune responses may be multifactorial, notably based on sex steroid concentrations, on transcriptional factors, and on incomplete inactivation of immunoregulatory genes on the second X chromosome in females(29, 30). Previous studies reported a relationship between ABO and rhesus blood groups, and COVID-19 susceptibility, suggesting that type O blood and rhesus-negative status may protect against severe COVID-19(31, 32). In our study, Rh-status was associated with faster decay of anti-S IgG titers over time, while no association was observed with ABO blood groups.

Although our study provides crucial data on the natural history of mild COVID-19, it is important to note that there are some limitations. Neutralization experiments were performed on a small subset of the cohort due to insufficient volume of remaining sera. However, the strong correlation between CMIA IgG levels and neutralizing titers observed in this study, and reported by the manufacturer as well as by other studies(15, 33, 34), allows an extrapolation of the results to the entire cohort. Assessment of reinfection was based on participant reports during visits, as no RT-PCR surveillance was planned in the study. Therefore, it cannot be excluded that the COVID-19 positive participants had unnoticed asymptomatic reinfection during follow-up. However, no COVID-19 positive HCW, except the case of reinfection, had a significant increase of both anti-S and anti-N levels during follow-up. Another limitation is the unbalanced sex distribution, with a female predominance, which reflects the sex distribution of the healthcare workers in our hospital. Nevertheless, the sex difference in immune response was observed by using univariate and multivariate analysis. Furthermore, we were not able to investigate the kinetics of memory B cells because of the lack of peripheral blood mononuclear cells. Finally, our results were obtained in participants with a median age of 39 years (IQR 30-51), hence we cannot exclude that older individuals may experience a different evolution of humoral response over time.

However, taken together our data demonstrate a long-term persistence of anti-S IgG titers that may protect convalescent COVID-19 patients against reinfection by variants D614G and B.1.1.7. By increasing the levels of cross-neutralizing antibodies, SARS-CoV-2 vaccine may strengthen their protection, especially against variants harboring antibody escape mutations like B1.351. Future work will help to determine whether vaccine-induced antibodies evolve in the same manner, and whether their kinetics differ between the sexes.

## Methods

### Study design and participants

We characterized SARS-CoV-2 antibody persistence in COVID-19 HCWs from Strasbourg University Hospital, France, up to 13 months after infection. Participants were recruited as follows (Figure 1): among 1,496 HCWs initially screened by SARS-CoV-2 serology between 6^th^ April and 7^th^ May 2020 in our institution, all participants with a COVID-19 history, proven either by serology at screening or by a previous RT-PCR, were recruited and followed at M1, M3-6, M7-9 and M11-13. In parallel, participants displaying negative serology without a history of positive RT-PCR for SARS-CoV-2 were recruited to evaluate the incidence of infection, and were followed by the same visit schedule as the COVID-19 positive cohort with M1 defined as one month post-inclusion. Participants completed a questionnaire at each visit in reference to sociodemographic characteristics, COVID-19 exposure, symptoms, virological findings and eventually vaccination.

### RT-PCR assay

RT-PCR for SARS-CoV-2 RNA detection was previously performed on nasopharyngeal swab samples at the time of diagnosis. All except six RT-PCR positive samples were analyzed in our laboratory with SARS-CoV-2 specific primers and probes targeting two regions on the viral RNA-dependent RNA polymerase (*RdRp*) gene (Institut Pasteur, Paris, France; WHO technical guidance). Ct values obtained in each sample were considered for statistical analyses.

### Serological assays

#### Screening assays

All sera were initially screened for SARS-CoV-2 antibodies using two commercial assays. The first is the Biosynex^®^ (COVID-19 BSS IgG/IgM) Lateral Flow Assay (LFA), which detects separately IgM and IgG directed against the Receptor Binding Domain (RBD) of the SARS-CoV-2 spike protein (S), with estimated overall sensitivity and specificity of 96% and 99% at 22 days since symptoms onset (DSO), respectively ^1^. The second assay used was the EDI™ Novel coronavirus COVID-19 IgG ELISA assay to detect anti-nucleocapsid protein (N) IgG at 22 DSO ^1^, which, in our hands, displayed a sensitivity of 81% and a specificity of 96%. The results rely on a ratio of specimen absorbance reported to the cut off (S/CO) value defined by the manufacturer.

#### Confirmation assay

All M1 sera associated with at least one positive result using the above-mentioned assays or with a history of positive SARS-CoV-2 RT-PCR were retrospectively analyzed with the Abbott Architect SARS-CoV-2 IgG Quant II assay (Abbott, Sligo, Ireland) to confirm the serological positive status and to measure the anti-S IgG titer, if allowed by remaining serum volume. Sera collected during follow-up were also analyzed with this commercial assay for the entire selected cohort of COVID-19 HCW to define the serological status at each timepoint. This assay is an automated chemiluminescence microparticle immunoassay (CMIA) that quantifies anti-RBD IgG, with 50 AU/mL as a positive cut-off and a maximal threshold of quantification of 40,000 AU/mL (80,000 AU/mL at 1:2 dilution). According to the manufacturer, this CMIA displays clinical sensitivity and specificity of 98.81% and 99.55% at 15 DSO, respectively. According to the manufacturer, antibody titers measured by this assay correlate, with a high probability (>95%), to neutralizing antibody titers assessed by plaque reduction assay on SARS-CoV-2 reference strain. This correlation was confirmed by previous studies(15, 33).

#### S-Fuse live-virus neutralization assay

A neutralizing assay was performed on a panel of 28 COVID-19+ participants, including 13 who received a single dose of COVID-19 vaccine. All sera were collected at M11-13. Live-virus neutralization was analyzed using the S-Fuse reporter cells, as previously reported (18). Briefly, S-Fuse reporter cells correspond to U2OS cells engineered to express-ACE2 and either GFP1–10 or GFP11. When mixed, these cells produce GFP upon syncytia formation which occurs during productive infection with SARS-CoV-2. Neutralization of infectious D614G, B.1.1.7 and B.1.351 variants was assessed for each serum using limiting-dilutions. Infection was quantified by measuring the number of GFP+ syncytia 18 hours after infection. The percentage of neutralization was calculated using the number of syncytia as the value with the following formula: 100 × (1 - (value with serum - value in ‘noninfected’)/(value in ‘no serum’ – value in ‘noninfected’)). Neutralizing activity of each serum was expressed as the half-maximal inhibitory concentration (IC_50_).

### Statistical analysis

Chi-squared test, Kruskal-Wallis rank sum test and Fisher’s exact test were conducted to identify any significant changes in categorical variables over time and between groups. Non-parametric Wilcoxon paired tests and Mann-Whiney tests were conducted to compare quantitative data over time or between groups, respectively. All tests were two-sided with an α level of 0.05. To model anti-S (log-transformed) and anti-N IgG titers over time, a triphasic decay was used, and the half-life (t_1/2_) of each decay phase was calculated.

To assess characteristics of patients with a faster/slower decay in anti-S and anti-N IgG titers, non-parametric tests were used for univariate analyses (Wilcoxon and Fisher’s exact tests). Variables achieving a *p* value <0.17 in the univariate analysis were entered into a multivariate linear regression model (with a backward stepwise method based on the likelihood ratio test). Multivariate analyses were performed with R software version 4.0.3 (R Foundation for Statistical Computing, Vienna, Austria). Factor effects in multivariate linear regression are given with a 95%CI. All other data were analyzed and represented using Graphpad Prism version 9.0.0. We used the Kaplan–Meier method to estimate the cumulative probability of SARS-CoV-2 infection or reinfection per group (COVID-19 negative or COVID-19 positive, respectively) and used the log-rank test to perform between-group comparisons. Time of exposition starts since the first negative serology for the COVID-19 negative HCW and since two months after initial SARS-CoV-2 infection (date of first symptoms or positive RT-PCR or first positive serology) for the COVID-19 positive group, as described previously(10). SARS-CoV-2 infection/reinfection was documented by RT-PCR or serological testing among COVID-19 positive and negative HCW, respectively. Vaccinated individuals were censored at the time of the first vaccine dose. We also calculated the incidence of SARS-CoV-2 infection/reinfection per month, according to at-risk days during the follow-up of COVID-19 positive and COVID-19 negative individuals.

### Study approval

This analysis was conducted on data from an on-going prospective, interventional, monocentric, longitudinal, cohort study enrolling healthcare workers from Strasbourg University Hospital (ClinicalTrials.gov Identifier: NCT04441684). The protocol was approved by the institutional review board of CPP Sud Méditerranée III. All participants provided a written informed consent.

## Data Availability

All data are available upon request to the corresponding author

## Author contributions

SFK, CSM, MG, YH and NM conceived and designed the study. CSM, JDS, NC, DR, MP AF, LKP and MG recruited participants. SB, NR, AS, LG, BP, and JML carried out LFA experiments. TB, DP and OS carried out S-Fuse neutralization experiments. FG, AV, MJW, MS and EL analyzed serological data. FG and PG performed statistical analyses. SFK, FG and PG wrote the manuscript with substantial input from all co-authors.

## Acknowledgments

We are grateful to all of the study participants who donated blood, the team of Cellule d’Appel, Pôle des Ressources Humaines who scheduled the planning of participants, the DRCI team including Eric Demonsant, Hélène Soavelo, Caroline Bouvrais and Evelyne Acacie who prepared the IRB protocol, the ECRF and the administrative part of the protocol, the CIC Inserm 1434 team, including Alexandre Bolle who coordinated recruitment of participants and Noelle Huber, Christine Toulouse, Sophie Wohlgemuth who collected samples, the team of Service de Medecine du Travail who managed the cohort, Carole Weber, Elodie Kleiber and Corinne Renaud for data management, and the technical staff of Pôle de Biologie and Laboratoire de Virologie including Anne Moncolin, Veronique Sohn, Axelle Grub, Nathalie Durand, Nadège Frey and Elise Suhr for management and distribution of samples. Manuscript edition and proofreading was provided by Dr. Kate Dunning (HUS). This work was supported by Strasbourg University Hospital (SeroCoV-HUS; PRI 7782), the Agence Nationale de la Recherche (ANR-18-CE17-0028), Laboratoire d’Excellence TRANSPLANTEX (ANR-11-LABX-0070_TRANSPLANTEX), and Institut National de la Santé et de la Recherche Médicale (UMR_S 1109).

## Notes

The authors have declared that no conflict of interest exists.

### Competing Interest Statement

The authors have declared no competing interest.

### Clinical Trial

ClinicalTrials.gov Identifier: NCT04441684

### Funding Statement

This work was supported by Strasbourg University Hospitals (SeroCoV-HUS; PRI 7782), the Agence Nationale de la Recherche (ANR-18-CE17-0028), Laboratoire d'Excellence TRANSPLANTEX (ANR-11-LABX-0070_TRANSPLANTEX), and Institut National de la Sante et de la Recherche Medicale (UMR_S 1109).

### Author Declarations

CPP Sud-Mediterranee 3, CHU de Nimes, Place du Professeur Robert Debre, 30029 Nimes Cedex 9 reference: 2020.04.15 bis_ 20.04.10.66856

### Summary of Updates

Correction of typos in the abstract and in the main text

